# Mass azithromycin distribution and cause-specific mortality among children aged 1-59 months: a secondary analysis of a cluster randomized controlled trial

**DOI:** 10.1101/2025.08.14.25333713

**Authors:** Ali Sié, Mamadou Ouattara, Mamadou Bountogo, Boubacar Coulibaly, Valentin Boudo, Thierry Ouedraogo, Elisabeth Gebreegziabher, Huiyu Hu, Elodie Lebas, Benjamin F. Arnold, Thomas M. Lietman, Catherine E. Oldenburg

**Affiliations:** Centre de Recherche en Santé de Nouna, Burkina Faso; Francis I Proctor Foundation, University of California, San Francisco, USA; Department of Epidemiology & Biostatistics, University of California, San Francisco, USA; Department of Ophthalmology, University of California, San Francisco, USA

## Abstract

Mass azithromycin distribution has been shown to reduce all-cause child mortality in several settings in the Sahel by 14-18%. A trial in Niger found that mass azithromycin distribution to children aged 1-59 months reduced cause-specific mortality due to malaria, dysentery, meningitis, and pneumonia. However, this study was done in the absence of seasonal malaria chemoprevention (SMC). Here, we assess the effect of mass azithromycin distribution on cause-specific child mortality in a setting receiving SMC. The Child Health with Azithromycin Treatment (CHAT) trial was a cluster randomized placebo-controlled trial of 341 communities in Nouna District, Burkina Faso. Eligible children (aged 1-59 months) received a single oral 20 mg/kg dose of azithromycin or matching placebo. Six rounds of distribution occurred over a 36-month period. An enumerative census was conducted during each twice-yearly distribution, during which vital status for all children in the community was collected. Verbal autopsy was performed to assess cause of death. Of 1,086 deaths recorded in the trial, verbal autopsy results were available for 992 (91%). The most common causes of death were infectious, including malaria (34%), diarrhea (24%), and pneumonia (9%). Children living in communities receiving azithromycin had significant reduction in malaria mortality (incidence rate ratio, IRR, 0.67, 95% confidence interval, CI, 0.50 to 0.90, P=0.008). Other infectious causes of mortality, including diarrhea and pneumonia, were lower in communities receiving azithromycin but were not statistically significantly different. Mass azithromycin distribution for child mortality has benefits in the context of SMC for reducing mortality, including for malaria mortality.

**Trial Registration:** ClinicalTrials.gov NCT03676764. Trial registered 17 September 2018.

## INTRODUCTION

Biannual mass azithromycin distribution has been shown in several cluster randomized trials to reduce all-cause child mortality when distributed periodically throughout the year.^1,2^ In the Niger site of MORDOR, a significant reduction in mortality due to malaria, dysentery, meningitis, and pneumonia was observed in azithromycin communities compared to placebo.^3^ MORDOR was done in the absence of seasonal malaria chemoprevention (SMC), a strategy that involves monthly distribution of sulfadoxine-pyramethamine (SP) and amodiaquine (AQ) to children aged 3-59 months during the high malaria transmission season in regions with highly seasonal malaria transmission.^4^ SMC is highly effective at reducing the incidence of clinical malaria, however the impact of SMC on mortality is less clear, with randomized controlled trials generally showing no effect of SMC on all-cause mortality.^5,6^ Following the results of MORDOR-Niger, debate ensued regarding whether mass azithromycin distribution would continue to be effective in other regions of the Sahel where SMC is routinely distributed, as malaria is one of the most common causes of mortality in the region.

The Child Health with Azithromycin Trial (CHAT) was a cluster randomized trial in Burkina Faso that evaluated mass azithromycin distribution in the presence of SMC.^7^ The trial found an 18% reduction in all-cause mortality in children aged 1-59 months, a point estimate that was identical to that observed in the Niger site of MORDOR.^1,2^ Despite the similarity in the reduction in all-cause mortality, the presence of SMC may change the distribution of cause of death, and azithromycin distribution may have different effects on cause-specific mortality in the presence of SMC compared to the absence SMC, particularly malaria. Here, we evaluated whether biannual mass azithromycin distribution reduced cause-specific mortality as assessed by verbal autopsy in the CHAT trial.

## METHODS

### Study Overview

Complete methods for the CHAT trial have been previously published.^2,7^ In brief, CHAT was a cluster randomized community trial in which 341 clusters in Nouna District, Burkina Faso were randomized to 6 rounds of twice-yearly mass distribution of azithromycin or matching placebo to children aged 1-59 months of age from August 2019 until February 2023. Clusters consisted of villages with < 2,000 inhabitants, or villages split into units with <2,000 population each for larger communities. The primary outcome for the trial was all-cause mortality and has been previously reported.^2^ 341 clusters were randomized to azithromycin or placebo; however, some clusters were lost over the course of the study due to security concerns in the study area. This was anticipated and accounted for in the trial’s statistical analysis plan. In total, 285 clusters contributed person-time to the analysis. The study was reviewed and approved by the Institutional Review Board at the University of California, San Francisco, the Comité d’Ethique pour la Recherche en Santé in Ouagadougou, Burkina Faso, and the Comité Technique d’Examen des Demandes d’Autorisation d’Essais Cliniques in Ouagadougou. Written informed consent was obtained from at least one guardian of each included child. The trial was registered at clinicaltrials.gov (NCT03676764).

### Study Setting

CHAT was conducted in Nouna District, Burkina Faso. Nouna is in northwestern Burkina Faso near the border with Mali in the Sahel. The region experiences seasonal rainfall from approximately July through October, which coincides with the high malaria transmission season. SMC with SP-AQ has been distributed monthly in July, August, September, and October in Nouna since 2014 via door-to-door distribution to children aged 3-59 months. SMC distribution therefore overlapped with azithromycin distribution from July-October in all years of the CHAT trial.

### Participants

Children were eligible for treatment as part of the trial if they weighed at least 3800 g at the time of the census and were between the ages of 1 and 59 months. Children were followed up to 65 months of age (6 months post last eligible treatment age) for vital status assessment.

### Randomization, Interventions, and Masking

Communities were randomized prior to the first census and treatment round to either biannual mass azithromycin distribution or placebo. Randomization was stratified by whether the community was within an existing Health and Demographic Surveillance Site (HDSS).^8^ Eligible children receive a single oral 20 mg/kg dose of azithromycin or equivalent volume of matching placebo (donated by Pfizer, Inc, New York, NY). All doses of study medication were directly observed. Study staff, investigators, participants, and caregivers were masked to the community’s randomized treatment allocation.

### Census

Every 6 months, a door-to-door enumerative census was performed in each study community. During the census, the head of each household was interviewed to determine the number of children under 5 years of age residing in the household, and the vital status of each child. Each census worker had a list of children in residence recorded during previous census rounds, and updated the vital status (alive, died, moved, unknown) for each known child, and added any new children who joined the household since the last census round. Study treatment (described below) was provided to all children aged 1-59 months at the time of the census.

### Cause of death assessment

A list of children who died per the census was generated during each census phase. After a mourning period of approximately 3 months, a verbal autopsy interview was attempted with the caregiver or head of household for each child that died using the 2016 World Health Organization Verbal Autopsy instrument for children aged 4 weeks to 11 years. Verbal autopsy results was assigned by inSilicoVA, an algorithm that assigns most likely cause of death, using R statistical software (R Foundation for Statistical Computing, Vienna, Austria).^9^ The primary cause of death as assigned by the algorithm was used as the cause of death in all analyses.

### Statistical analysis

The overall distribution of causes of death was compared between arms using multivariate analysis of variance (MANOVA). We estimated incidence rate ratios (IRR) and incidence rate differences (IRD) and corresponding 95% confidence intervals (CI) for each cause of death separately using Poisson regression models with robust standard errors. All analyses were conducted at the cluster level (the level of randomization). As a secondary analysis, we assessed the effect of azithromycin on all infectious cases of mortality using similar methods. All analyses were conducted in Stata version 17.0 (StataCorp, College Station, TX). All statistical tests were two-sided with a P-value of < 0.05 considered statistically significant.

## RESULTS

Of 341 communities originally randomized as part of the trial, 146 in the azithromycin group and 139 in the placebo group contributed person-time to the trial and were included in the analysis (**Figure 1**). Communities were treated from August 2019 through February 2023. Of the 341 originally randomized communities, 56 did not contribute person-time due to security concerns that arose during the conduct of the trial.^2^ During the baseline census, the distribution of age and sex was similar between children residing in communities in the azithromycin and placebo groups (**Table 1**). The primary outcome, all-cause mortality, has previously been reported.^2^ The primary outcome included 498 deaths in the azithromycin group and 588 in the placebo group. Of these, verbal autopsies were available for 456 (91.6%) deaths in the azithromycin group and 536 (91.2%) in the placebo group.

**Figure 1.**
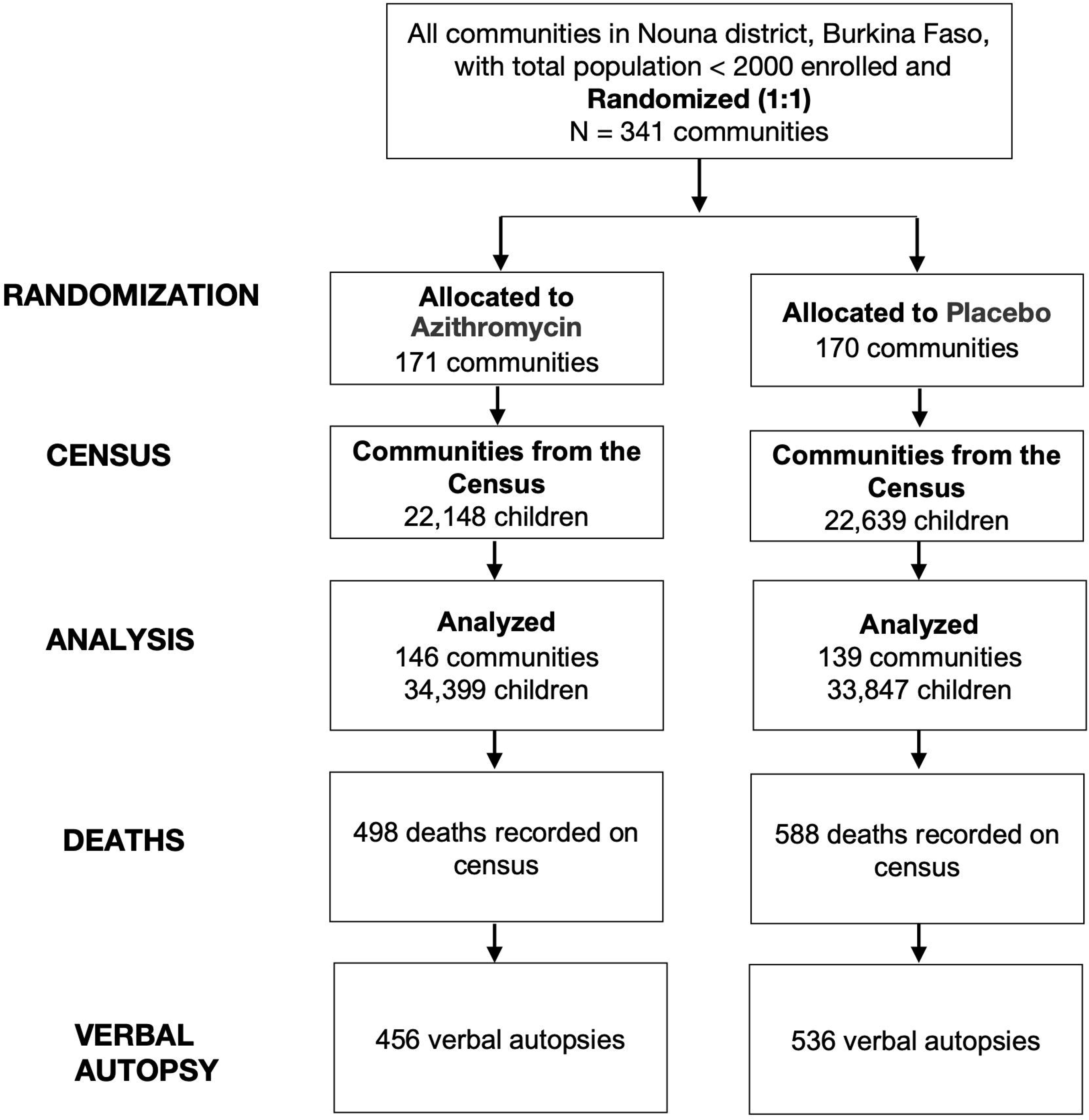
CONSORT diagram of study participation. **Figure 1 Alt Text.** Flow diagram showing number of communities randomized, allocated to azithromycin and placebo, number of children included in analyses, and number of deaths recorded on the census and with verbal autopsy data available.

**Table 1.** Community characteristics at the baseline census. **Table 1 Alt Text.** Baseline demographics table showing distribution of baseline characteristics at the community level between the azithromycin and placebo groups.

|  | **Azithromycin group** | **Placebo group** |
| --- | --- | --- |
| Number of communities | 144 | 140 |
| Number of children | 22,148 | 22,639 |
| Age group |  |  |
| 1–11 months | 3606 (16.3%) | 3580 (15.8%) |
| 12–23 months | 4575 (20.7%) | 4709 (20.8%) |
| 24–59 months | 13967 (63.1%) | 14,350 (63.4%) |
| Sex |  |  |
| Male | 11,265 (50.9%) | 11,390 (50.3%) |
| Female | 10,883 (49.1%) | 11,249 (49.7%) |

The overall distribution of deaths did not differ between azithromycin and placebo communities (*P*=0.21). Infectious causes were the most common for all calendar months of the year (N=866, 80%, over entire study period), with an increase in the number of malaria deaths in the high malaria transmission season (**Figure 2**). The most common cause of death per verbal autopsy was malaria (**Table 2**). There were 127 malaria-associated deaths in the azithromycin group (incidence rate 2.3 malaria deaths per 1,000 person-years) and 181 malaria deaths in the placebo group (3.4 malaria deaths per 1,000 person-years), corresponding to a 33% reduction in malaria mortality in azithromycin-treated communities compared to placebo (IRR 0.67, 95% confidence interval, CI, 0.50 to 0.90, **Figure 3**). Diarrhea was the second-most common cause of mortality, with 98 (1.8 diarrhea deaths per 1,000 person-years) in the azithromycin group and 118 (2.2 diarrhea deaths per 1,000 person-years) in the placebo group (IRR 0.79, 95% CI 0.54 to 1.16), followed by pneumonia and meningitis (**Table 2**). An analysis pooling all infectious causes of mortality into a single group (all-cause infectious mortality) found an infectious mortality rate of 7.1 deaths per 1,000 person-years in the azithromycin group and 8.8 deaths per 1,000 person-years in the placebo group, consistent with an approximately 20% reduction in infectious mortality in communities receiving azithromycin (IRR 0.80, 95% CI 0.64 to 1.01). The most common non-infectious cause of mortality was injury, and there was no evidence of a difference in injury-related mortality between groups (**Table 2**; **Figure 3**).

**Figure 2.**
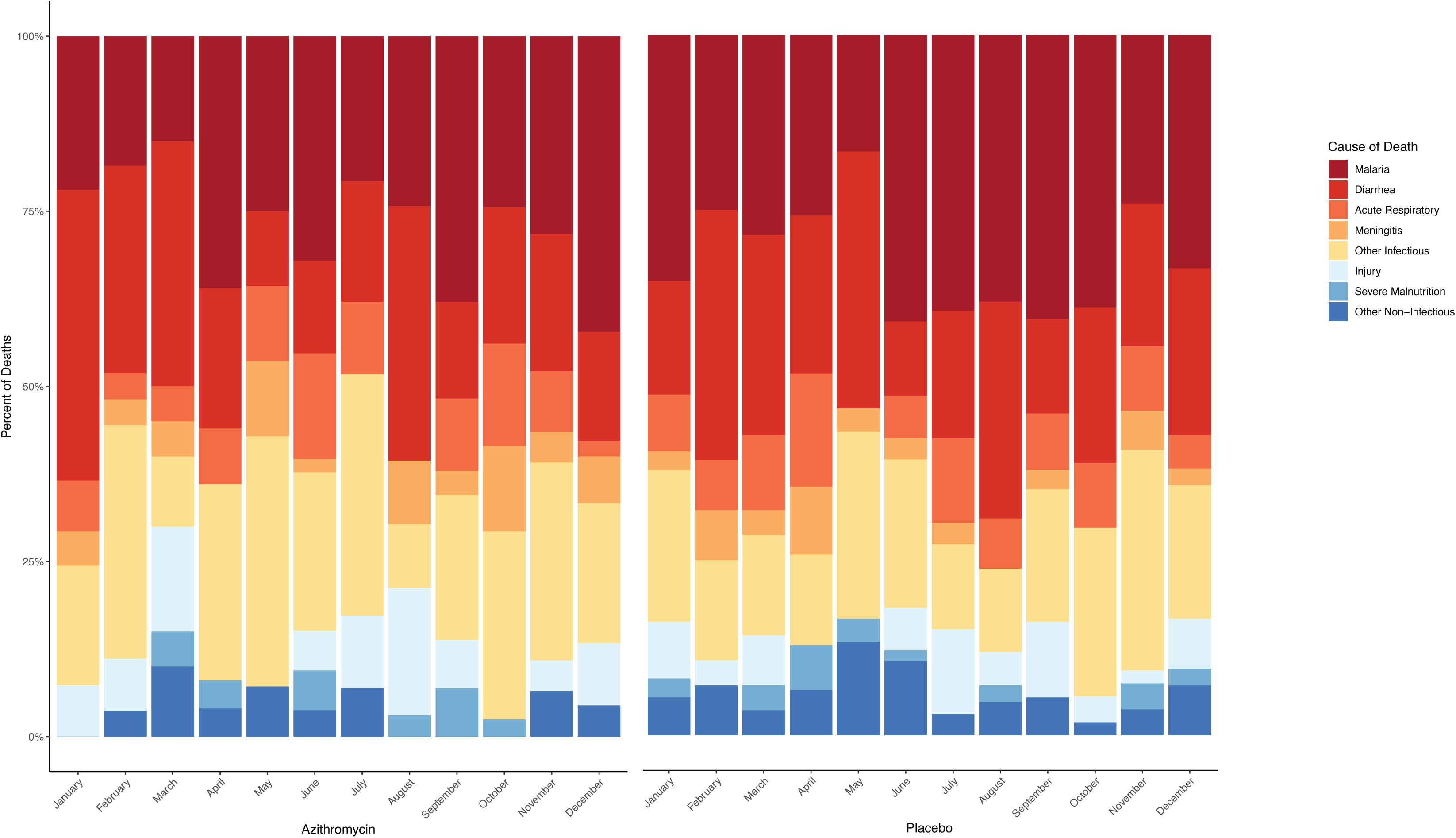
Cause of death as assigned by verbal autopsy in children living in communities randomized to azithromycin (left) and placebo (right) distribution by month of death. **Figure 2 Alt Text.** Stacked bar chart showing the distribution of cause of death in the azithromycin and placebo groups, with cause of death color coded. The figure shows that the most common cause of mortality as assigned by verbal autopsy is malaria, followed by diarrhea.

**Figure 3.**
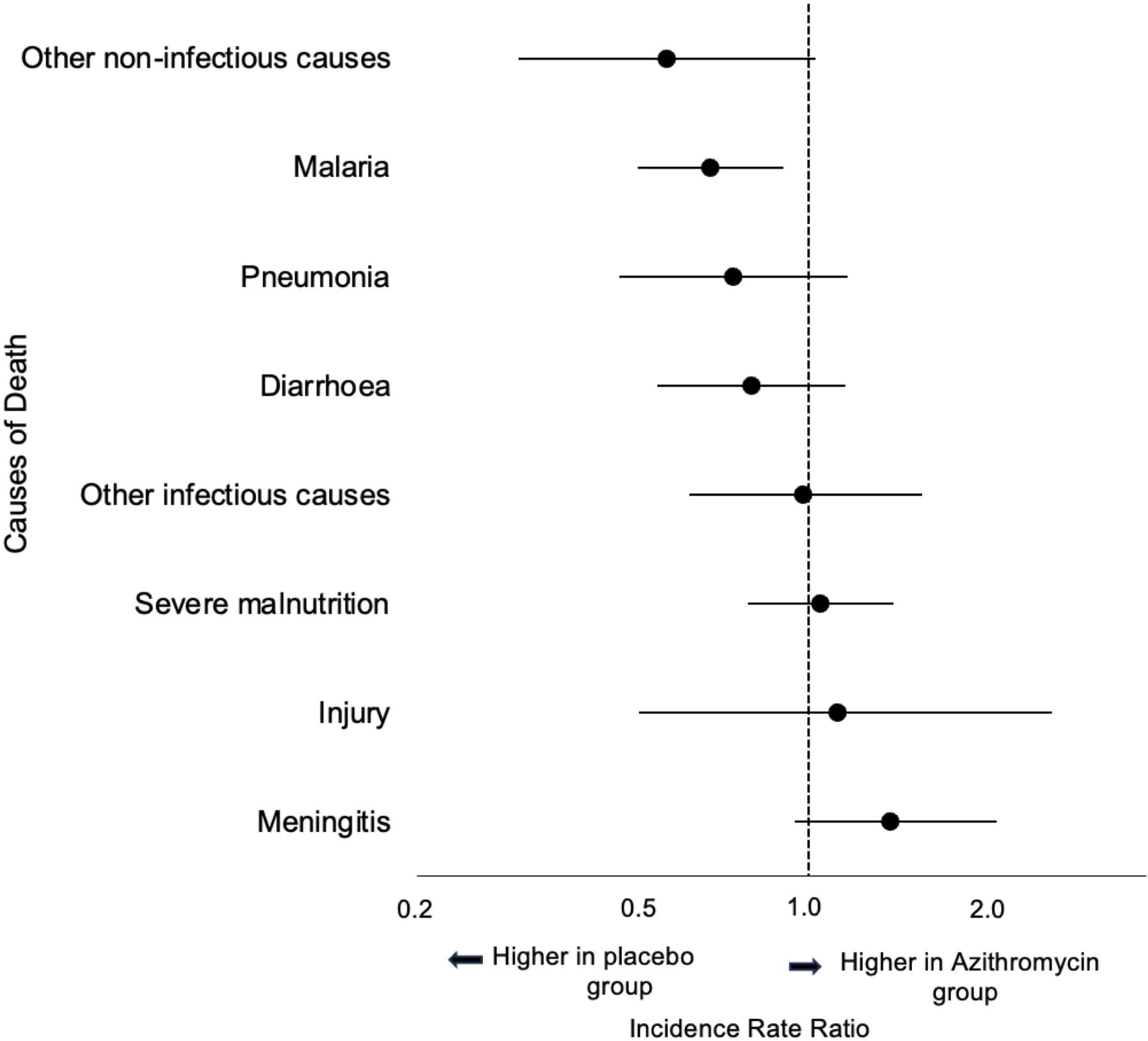
Effect of azithromycin compared to placebo on cause-specific mortality. Point estimates are incidence rate ratios and bars are 95% confidence intervals. **Figure 3 Alt Text.** Forest plot showing point estimates and 95% confidence intervals for each verbal autopsy-assigned cause of death in the placebo compared to azithromycin groups.

**Table 2.**
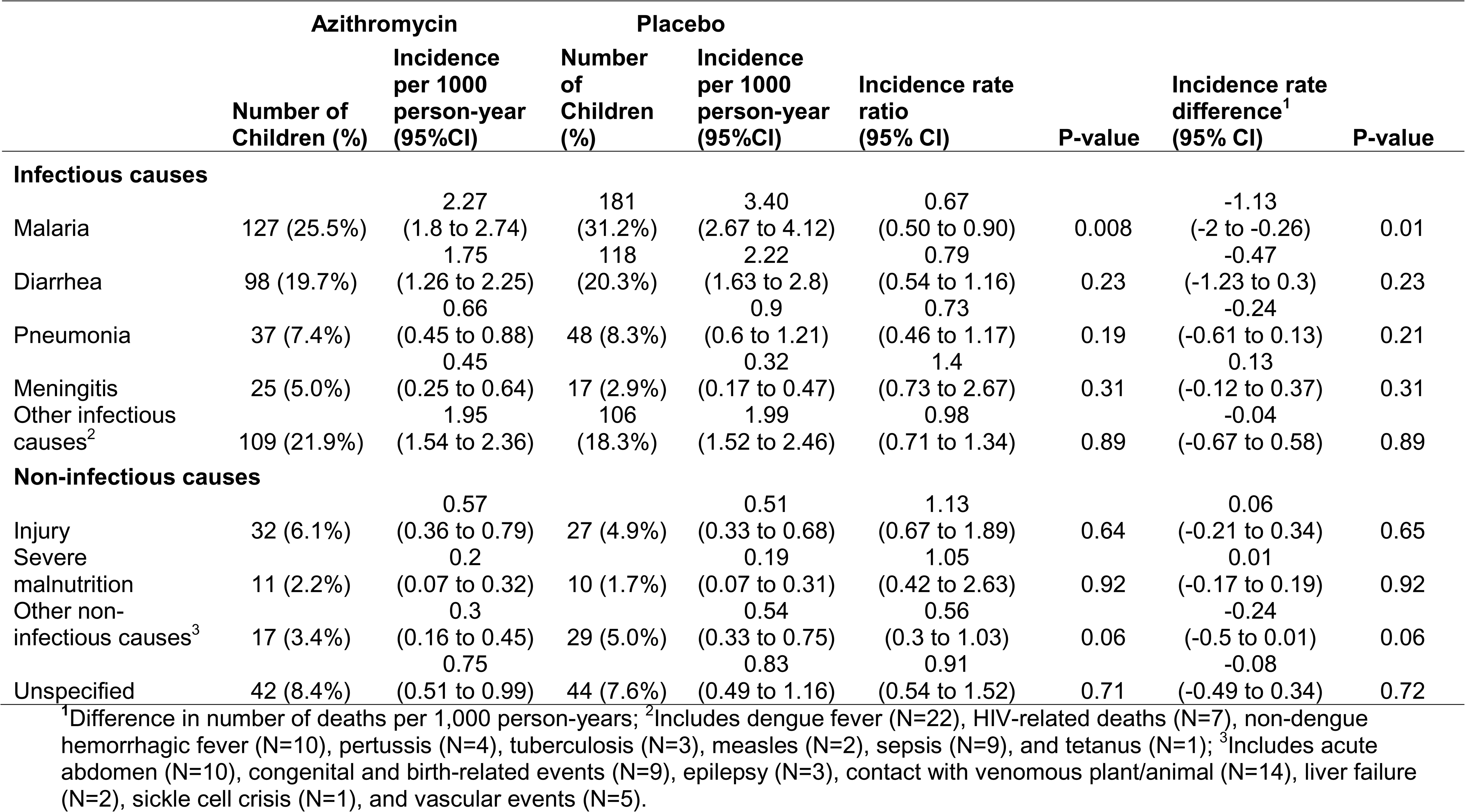
Primary cause of death as assigned by verbal autopsy in communities randomized to azithromycin and placebo. **Table 2 Alt Text.** Table showing incidence rates per 1000 person-years for each verbal autopsy assigned cause of death along with incidence rate ratios and differences comparing each cause-specific cause of death in the azithromycin and placebo groups.

## DISCUSSION

In this secondary analysis of verbal autopsy data from a cluster randomized trial of biannual mass azithromycin distribution compared to placebo for prevention of mortality in Burkina Faso, we found evidence of a reduction in malaria mortality as determined by verbal autopsy in children living in azithromycin compared to placebo communities. The primary analysis of the trial found an 18% reduction in all-cause child mortality in communities receiving azithromycin compared to placebo, a result consistent with the Niger site of the MORDOR trial.^1,2^ The CHAT trial was performed in the presence of SMC; communities in CHAT have received monthly SMC from July through October since 2014. In contrast, SMC distribution had not yet begun in the districts in Niger in which the original MORDOR trial was conducted. A previous household randomized controlled trial comparing SMC distribution with and without azithromycin found no additional benefit of adding azithromycin to SMC treatment regimens.^10^ This led to a hypothesis that there may be no additional benefit of mass azithromycin when SMC is also being distributed, given that malaria is a major cause of mortality in the study area and that SMC reduces malaria mortality and sulfadoxine may have non-malarial activity against pathogens that contribute to child mortality.^5,11^ In the present analysis, the malaria mortality was more common during the SMC season and malaria mortality was significantly reduced in communities receiving azithromycin compared to placebo.

Azithromycin has weak antimalarial properties as it targets the plasmodial apicoplast.^12,13^ Previous studies have found limited evidence of an effect of mass azithromycin distribution for reduction in parasitemia and serologic responses to MSP-1, a surrogate for malaria incidence.^14–21^ One study suggested a short-term reduction in malaria parasitemia following mass azithromycin distribution for trachoma^17^, and other studies have documented small reductions in parasitemia in communities receiving azithromycin compared to placebo for prevention of child mortality and in communities receiving biannual compared to annual azithromycin for trachoma.^20,21^ However, other studies have found no effect of azithromycin on parasitemia^18^, and individually randomized studies of single dose azithromycin have not shown an effect of azithromycin on short- or longer-term parasitemia prevalence.^22,23^ Analysis of verbal autopsy data showed a 22% reduction in malaria mortality in the MORDOR trial^3^, consistent with findings from the present study. An analysis of the CHAMPS study documented a high prevalence of bacterial co-infection in children who died from malaria.^24^ If direct effects of azithromycin on malaria are insufficient to reduce malaria mortality, it is possible that azithromycin distribution could reduce bacterial co-infections that may increase malaria-related mortality.

Malaria was the most commonly assigned cause of death per the verbal autopsy. Pneumonia was less common than expected based on cause-specific healthcare visits for children under 5, for which pneumonia is the most common^25,26^, and cause of death estimates from sub-Saharan Africa.^27^ Deaths in the study area were identified via biannual census, often occurring outside of the health system and no physician-assigned cause of death information or diagnostic information was available. Verbal autopsy has generally been shown to have low sensitivity but relatively higher specificity for assigning malaria mortality.^24,28–30^ A previous analysis of CHAMPS data showed 29% sensitivity and 87% specificity of the inSilicoVA algorithm for assigning malaria as a cause of death versus minimally invasive tissue sampling.^24^ Although we did not have a gold standard method of assigning cause of death in the present study, if the CHAMPS are results are generalizable to the CHAT study, this would suggest that malaria mortality would be undercalled rather than overcalled. If misclassification of cause of death was non-differential with respect to randomized treatment assignment, this would likely bias result towards the null.

In addition to limitations due to potential misclassification due to the use of verbal autopsy and absence of a gold standard method of assigning cause of death, there are several limitations to consider in this analysis. First, even in high mortality settings, mortality is a rare outcome.

Confidence intervals were wide for all causes of mortality, and especially for those that were less common. Although the finding is consistent with the MORDOR study, it is possible that the reduction in malaria mortality was due to chance. For example, while the confidence interval included 1, there appeared to be a reduction in mortality due to non-infectious causes that were not due to accident (e.g., snake bite) in azithromycin-treated communities. However, this is most likely a chance finding, due to the lack of biological plausibility for an association between azithromycin distribution and non-infectious causes of mortality. Non-malarial infectious causes of mortality, including pneumonia and diarrhea, were non-statistically significantly lower in azithromycin communities, but confidence intervals were wide due to low statistical power. An analysis considering all infectious causes of mortality found approximately 20% reduced risk of infectious mortality in communities receiving azithromycin compared to placebo, although the confidence interval narrowly included the null. Second, verbal autopsies were not available for all deaths. Verbal autopsies are performed several months after the death occurs, to honor the mourning period. Families may be untraceable after this period. The percentage of deaths for which a verbal autopsy was available was similar between the two study arms, and missingness was not differential by arm. Third, results may not be generalizable to settings with a different distribution of causes of mortality. Although results of the effect of biannual azithromycin distribution on mortality have been broadly consistent in studies conducted in the Sahel (e.g., the Niger site of MORDOR and the CHAT study), these results may not be generalizable outside of the Sahel.^31,32^

In this analysis of cause-specific mortality, we found a reduction in verbal autopsy-assigned malaria mortality in communities receiving biannual mass azithromycin distribution to children aged 1-59 months compared to placebo. This finding was consistent with previous studies of mass azithromycin distribution for prevention of child mortality in the Sahel and suggests that there are likely additional beneficial effects of azithromycin distribution even in the presence of SMC. Further studies with gold standard malaria and malaria mortality assessments are warranted.

## Data Availability

All data produced in the present study are available upon reasonable request to the authors

## AUTHOR CONTACT INFORMATION

Ali Sié: Centre de Recherche en Santé de Nouna, Burkina Faso;

Mamadou Ouattara: Centre de Recherche en Santé de Nouna, Burkina Faso;

Mamadou Bountogo: Centre de Recherche en Santé de Nouna, Burkina Faso;

Boubacar Coulibaly: Centre de Recherche en Santé de Nouna, Burkina Faso;

Valentin Boudo: Centre de Recherche en Santé de Nouna, Burkina Faso;

Thierry Ouedraogo: Centre de Recherche en Santé de Nouna, Burkina Faso;

Elisabeth Gebreegziabher: University of California, San Francisco;

Huiyu Hu: University of California, San Francisco;

Elodie Lebas: University of California, San Francisco;

Benjamin F. Arnold: University of California, San Francisco;

Thomas M. Lietman: University of California, San Francisco;

Catherine E. Oldenburg: University of California, San Francisco;

